# Comparison of anxiety and satisfaction levels in patients undergoing digital versus conventional dental impressions

**DOI:** 10.64898/2026.02.05.26345703

**Authors:** María Emilia Farfán, Ana Paula Pinzón, Marcelo Armijos Briones

**Affiliations:** School of Dentistry, Universidad de Especialidades Espíritu Santo, Samborondón, Ecuador

**Keywords:** Dental Impression Technique, Dental Impression Materials, Dental Anxiety, Patient Satisfaction, Imaging, Three-Dimensional, Computer-Aided Design

## Abstract

Dental impressions are a routine component of prosthodontic care, yet the patient experience may vary depending on the technique used. This study compared dental anxiety and satisfaction among patients undergoing digital versus conventional impressions in a postgraduate clinical setting at the Universidad de Especialidades Espíritu Santo. A total of 85 adult patients were included: 44 received conventional impressions and 41 received digital impressions. Dental anxiety was assessed using the Modified Dental Anxiety Scale (MDAS) before the procedure, and satisfaction was evaluated using a Visual Analog Scale (VAS) immediately after the procedure. Anxiety scores did not differ significantly between groups (p = 0.232). However, patients in the digital group reported significantly greater satisfaction than those in the conventional group (p < 0.001). These findings suggest an association between the use of digital impression techniques and higher levels of patient satisfaction, while no significant association was observed between impression technique and dental anxiety.

## Introduction

Dental impressions, as emphasized by numerous studies, continue to be a cornerstone of modern dental practice, especially in prosthetics, orthodontics, and oral rehabilitation [1]. These procedures allow for accurate replication of intraoral structures, which is essential for the fabrication of prostheses and other oral devices [2]. Traditionally, materials such as alginate and silicone have been used, requiring patient cooperation and tolerance during potentially uncomfortable procedures [3].

However, conventional impression techniques are not free from limitations, as they have been shown to cause physical discomfort, gag reflex, and breathing difficulties in a significant portion of patients [4]. One clinical study comparing alginate impressions with digital scanning reported that conventional methods resulted in notably higher levels of discomfort, including nausea, gagging, and general intolerance during the procedure [4]. Further, patients with an exaggerated gag reflex and elevated dental anxiety were found to be more prone to anticipatory nausea and treatment avoidance, underscoring the emotional burden imposed by these techniques [5]. Additionally, the use of bulky trays and extended intraoral working time associated with conventional impressions has been linked to heightened sensations of suffocation and stress, resulting in significantly higher anxiety levels when compared to impression methods adapted for greater patient comfort [6]. These adverse effects may reduce patient satisfaction and lead to treatment avoidance in more severe cases [7].

The emergence of intraoral scanning has revolutionized dental impression-taking [8]. Digital impressions offer a highly accurate, non-invasive alternative that eliminates the need for trays and impression materials [9]. These methods provide superior precision, real-time visualization, and improved communication between clinician and dental laboratory; moreover, digital techniques tend to reduce chair time and offer a more patient-friendly experience [10–12].

Recent literature has shown that digital impressions exhibit superior trueness and reproducibility compared to conventional methods [13]. In oral rehabilitation, where treatment quality is directly linked to patient cooperation and adaptation, digital workflows are particularly advantageous, as they not only improve procedural efficiency but also foster greater trust and acceptance among patients [14]. According to findings on dental anxiety prevalence, most adults attending public dental clinics report moderate to high levels of dental anxiety, with 31.8% postponing dental visits, 59.1% of whom do so because of fear [15]. In this context, digital techniques may provide a less stressful alternative, achieving up to 94% clinically viable dental segmentation without the need for additional corrections [16], further supported by non-invasive anxiety-reduction strategies such as music therapy, with music therapy showing significant effects in reducing anxiety, particularly in children [17]. Elevated anxiety levels can impair communication, reduce pain tolerance, and limit cooperation during procedures, addressing patient anxiety becomes a critical factor that may ultimately determine the success or failure of the treatment, regardless of the technical accuracy achieved.

Dental anxiety remains a critical concern. Globally, approximately 15% of adults report significant levels of dental fear, often triggered by a perceived lack of control, fear of pain, and previous negative experiences. These emotional factors can influence treatment avoidance and overall oral health outcomes [18,19]. Conventional impressions, often associated with invasive sensations, may aggravate this anxiety [20]. In contrast, digital impressions have been shown to lower anxiety levels and increase comfort, especially in patients with previous negative experiences [21–23].

Recent literature highlights that patient satisfaction is not solely determined by the technique itself but also by the environment, operator skills, and psychological factors [24,25]. While most patients report greater comfort and satisfaction with digital impressions compared to conventional techniques, some studies indicate that this preference is not universal [26,27]. Although digital impressions are generally associated with reduced gag reflex, shorter procedure time, and greater patient comfort, variations still exist depending on the dental condition and the complexity of the scan, this review highlighted that in some clinical settings, particularly those involving extended edentulous spans or mobile soft tissues, conventional impressions may still be perceived as more reliable or suitable by certain patients, especially those unfamiliar with digital workflows [26]. Similarly, a systematic review by Ahmed et al. [27] concluded that while digital impressions show significant advantages in terms of operator convenience and patient preference, not all patients experience a substantial improvement in comfort. Their findings emphasized that individual factors, such as prior dental experiences, anxiety levels, and personal expectations, play a critical role in shaping patient satisfaction; it is evident that despite the many advantages of digital impressions, patient perceptions remain multifactorial and context dependent.

Given the increasing emphasis on patient-centered care, it is essential to explore these emotional and experiential dimensions. Academic clinical environments, such as university postgraduate clinics, represent unique contexts for assessing both clinical effectiveness and patient satisfaction [28,29]. Tools like the Modified Dental Anxiety Scale (MDAS) and the Visual Analog Scale (VAS) allow for reliable measurement of psychological and experiential outcomes [30,31]. In contrast, digital impressions have demonstrated multiple advantages: less invasiveness, real-time visualization, greater precision, and improved patient comfort [10,14]. These benefits can translate to enhanced patient trust and satisfaction, better clinical efficiency, and higher long-term treatment success [32,33].

Despite numerous studies comparing digital and conventional impressions, gaps in the literature still exist [6,9]. Most studies focus on technical accuracy and immediate patient satisfaction, but few have evaluated the long-term impact of these techniques on dental anxiety or treatment perception. For instance, research on how previous experiences with digital impressions may influence the patient’s future expectations is limited. Additionally, there is a lack of longitudinal studies that follow patients over an extended period after treatment to evaluate the durability and effectiveness of prostheses based on digital impressions. This lack of long-term studies limits the understanding of how digital impressions affect not only the patient’s immediate experience but also their long-term quality of life and overall perception of dentistry.

Consequently, the lack of research in these areas suggests the need for future studies exploring the lasting impact of impression techniques on patient anxiety and satisfaction, as well as their relationship to treatment adherence [7].

This study aims to determine and compare the levels of dental anxiety and patient satisfaction in individuals undergoing digital and conventional impressions in a postgraduate clinical setting. Using the Modified Dental Anxiety Scale (MDAS) and the Visual Analog Scale (VAS), the research seeks to explore how each technique affects patient comfort and emotional response. The findings are expected to provide valuable insights to guide clinicians and educators in making patient-centered decisions and improving the quality of prosthodontic care.

## Materials and methods

### Study Design

A cross-sectional, observational, and comparative study was conducted to evaluate the levels of anxiety and satisfaction among patients undergoing digital versus conventional dental impressions. The study was carried out at the Postgraduate Clinic of the Oral Rehabilitation and Implant-Assisted Prosthodontics Specialty at the Universidad de Especialidades Espíritu Santo, located in Samborondón, Ecuador. Data collection took place between May 1 and June 30, 2025.

The protocol was reviewed and approved by the Ethics Committee of the Universidad de Especialidades Espíritu Santo under approval code C-UEES-25-01 (see S1 File).

### Participants

The study sample consisted of adult patients (≥18 years) who attended the clinic during the established period and required preliminary or functional dental impressions as part of their prosthetic or restorative treatment plan (fixed prosthesis, removable partial denture, complete denture, implant-supported prosthesis, or indirect restorations). A consecutive sampling method was used, in which all patients who met the inclusion criteria and attended during the study period were incorporated in order of arrival, without omissions, until the data collection period was completed.

We used consecutive sampling over a fixed two-month period. All eligible adult patients attending the Oral Rehabilitation Postgraduate Clinic during the study period were invited to participate while waiting for the appointment. Those who agreed signed informed consent and were enrolled consecutively.

Group allocation was not determined by the research team. Participants were classified into the “digital” or “conventional” group according to the impression technique planned by the treating postgraduate clinician as part of routine care. The choice of technique was based on clinical considerations and feasibility, including the patient’s ability to cover the additional cost of digital scanning.

### Inclusion Criteria

- Patients who required preliminary or functional dental impressions for a fixed prosthesis, removable partial denture, complete denture, implant-supported prosthesis, or indirect restoration, either of the following:

- Conventional impressions performed using alginate, silicone, stock trays, or custom full trays, depending on the clinical case.
- Digital impressions, obtained through an intraoral scanner.
- Age 18 years or older.
- Signed informed consent and compliance with the completion of the anxiety and satisfaction questionnaires.

### Exclusion Criteria

- Patients who decline to participate or fail to complete the questionnaires.
- Patients with cognitive impairments that prevent effective participation.
- Patients who have been diagnosed with anxiety.

### Variables and Instruments

The primary variables analyzed in this study were dental anxiety and patient satisfaction:

Dental anxiety (MDAS) was assessed immediately before the impression procedure to capture anticipatory anxiety and to avoid contamination by the procedural experience itself. Post-procedure assessment could be influenced by relief, discomfort, or immediate procedural outcomes, potentially biasing anxiety scores. Patient satisfaction was assessed immediately after the procedure using the VAS-based questionnaire, as satisfaction requires the participant to have experienced the impression process.

#### Dental anxiety

Participants’ anxiety levels were assessed prior to undergoing the dental impression procedure using the Modified Dental Anxiety Scale (MDAS) (see S2 File). This validated scale consists of five items that evaluate different aspects of anxiety related to dental care, with responses ranging from 1 (relaxed, not anxious at all) to 5 (extremely anxious). Total scores range from 5 to 25, with less than 9 indicating mild or no anxiety, 9–12 indicating moderate anxiety, 13–14 indicating high anxiety, and 15 or more indicating severe anxiety. Higher scores indicate greater anxiety [34,35].

Anxiety was assessed prior to the dental impression procedure to reflect participants’ pre-procedural anxiety. Measuring anxiety after the procedure could have influenced the results due to discomfort or the experience of the impression itself.

#### Patient satisfaction

Patient satisfaction was assessed immediately after completing the dental impression procedure using the Visual Analog Scale (VAS), validated by researchers at The Ohio State University (see S3 File). The survey consisted of seven questions addressing comfort, procedure duration, gag sensation, and overall satisfaction. Each item was rated from 1 (strongly agree) to 5 (strongly disagree), with total scores ranging from 7 to 35. Lower scores indicate higher satisfaction [33].

Satisfaction was assessed after completion of the dental impression procedure so that participants could evaluate their experience based on the procedure they had just undergone. This timing allowed responses to reflect perceptions of comfort, duration, and overall experience, which cannot be accurately assessed before the impression is completed.

For this study, the data were collected through a survey created in Google Forms. The questionnaires were completed inside the Postgraduate Oral Rehabilitation Clinic, in the same space where the impression procedures were carried out. Before answering the questions, participants first went through the informed consent, which appeared as the first section of the form, they could only move on to the survey after selecting “I agree.” Both surveys were short 7 questions for anxiety and 5 for satisfaction so filling them out took around 2 to 4 minutes. All responses were recorded directly in Google Forms and later transferred to an Excel spreadsheet for subsequent analysis.

### Statistical Analysis

Normality was assessed using the Kolmogorov-Smirnov test and visual inspection of distributions. Given the non-normal distribution and ordinal nature of the scales, non-parametric tests were applied. Due to this, the Mann–Whitney U test was used to determine differences in anxiety and satisfaction scores between the two impression groups. To analyze differences among participants with respect to sex and marital status, the Chi-square test was applied. For the variables household income and age, the mean and standard deviation were used for descriptive presentation. A p-value < 0.05 was considered statistically significant.

## Results

A total of 85 patients were included in the study. Of these, 44 individuals (51.8%) underwent conventional dental impressions, while 41 (48.2%) received digital impressions. According to the data presented in **Table 1** the sample was evenly distributed between groups, allowing for balanced statistical comparisons. There were no statistically significant differences between the conventional and digital impression groups in terms of age, monthly income, sex, or marital status (all p > 0.05). Although baseline sociodemographic variables were similar, unmeasured clinical and economic factors related to treatment indication and affordability may still differ between groups.

**Table 1.**
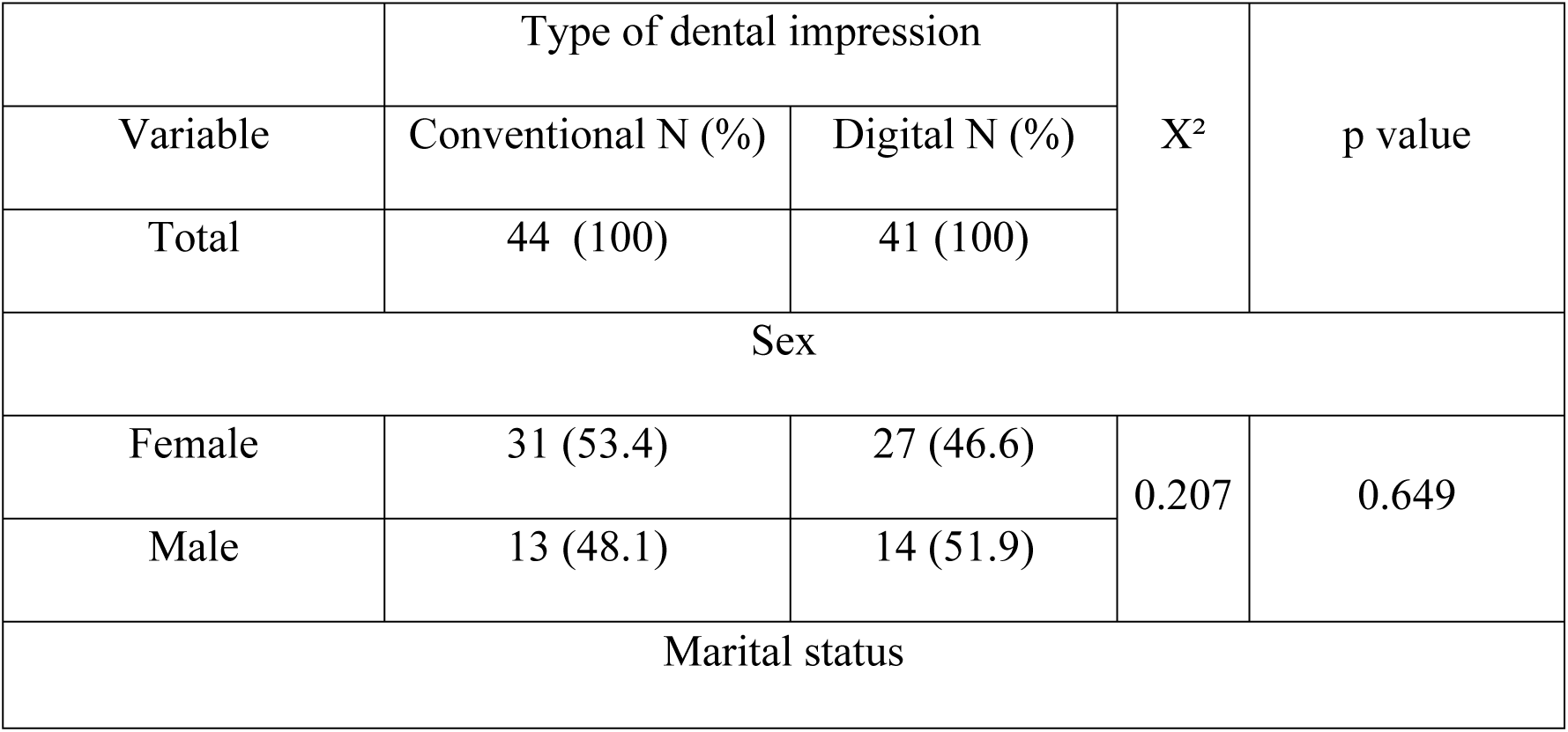
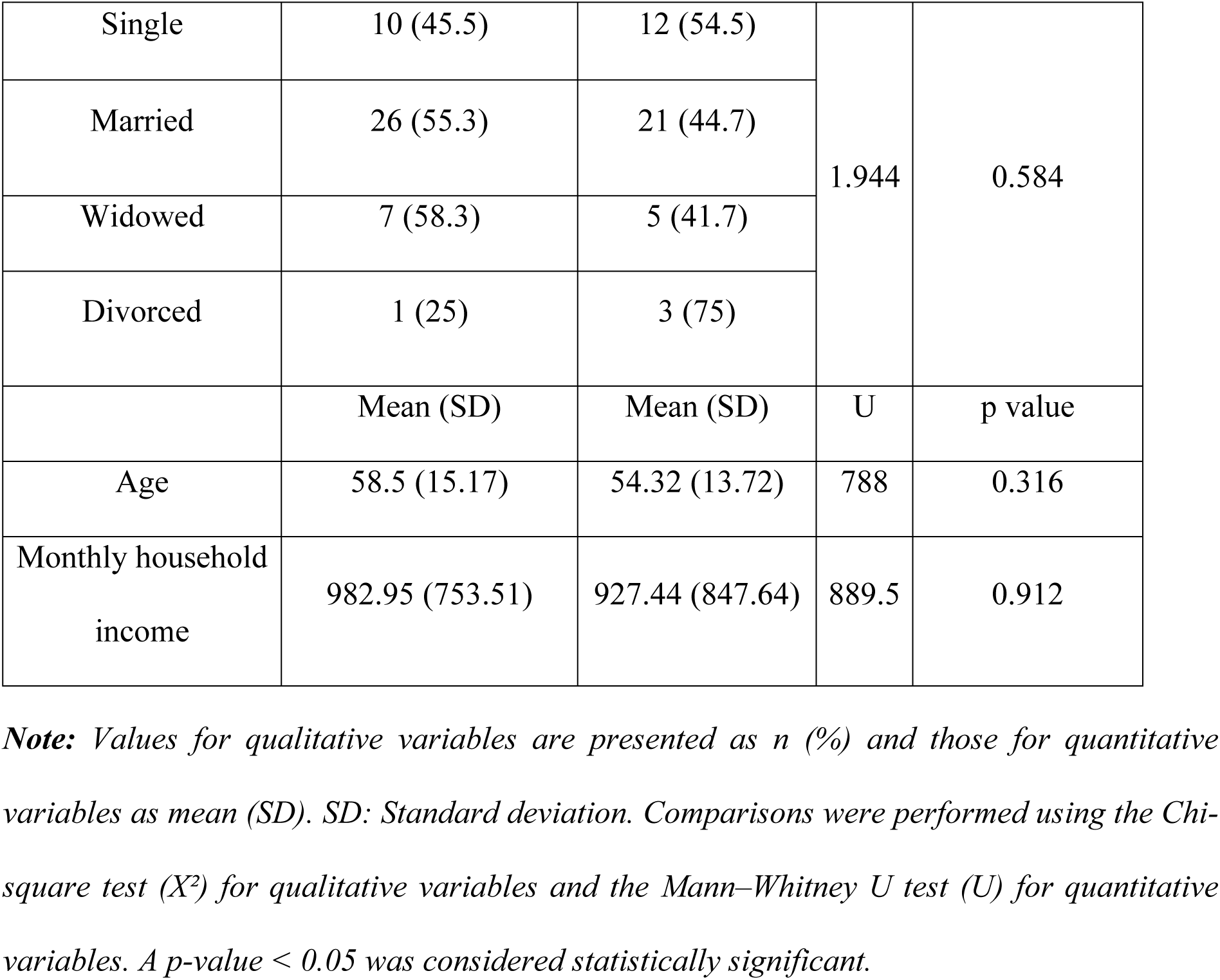
Sociodemographic and economic characteristics by type of dental impression.

|  | Type of dental impression |  | X<sup>2</sup> | p value |
| --- | --- | --- | --- | --- |
| Variable | Conventional N (%) | Digital N (%) |  |  |
| Total | 44 (100) | 41 (100) |  |  |
| Sex |  |  |  |  |
| Female | 31 (53.4) | 27 (46.6) | 0.207 | 0.649 |
| Male | 13 (48.1) | 14 (51.9) |  |  |
| Marital status |  |  |  |  |
| Single | 10 (45.5) | 12 (54.5) | 1.944 | 0.584 |
| Married | 26 (55.3) | 21 (44.7) |  |  |
| Widowed | 7 (58.3) | 5 (41.7) |  |  |
| Divorced | 1 (25) | 3 (75) |  |  |
|  | Mean (SD) | Mean (SD) | U | p value |
| Age | 58.5 (15.17) | 54.32 (13.72) | 788 | 0.316 |
| Monthly household income | 982.95 (753.51) | 927.44 (847.64) | 889.5 | 0.912 |

*Note: Values for qualitative variables are presented as n (%) and those for quantitative variables as mean (SD). SD: Standard deviation. Comparisons were performed using the Chi-square test ( $X^2$ ) for qualitative variables and the Mann–Whitney U test (U) for quantitative variables. A p-value < 0.05 was considered statistically significant.*

The participants included in this study were evaluated with the Modified Dental Anxiety Scale (MDAS) before the impression-taking procedure. Patients in the conventional group obtained a mean score of 8.91 (standard deviation = 3.326), while the digital group had a mean score of 8.24 (standard deviation = 3.239). As shown in **Table 2**, the Chi-square and Mann–Whitney U tests revealed no statistically significant differences between the groups (U = 767.5; p = 0.232).

**Table 2.** Comparison of anxiety levels between conventional and digital impressions.

|  | Mean (M) | Standard deviation (SD) | U | p value |
| --- | --- | --- | --- | --- |
| Conventional | 8.91 | 3.326 | 767.5 | 0.232 |
| Digital | 8.24 | 3.239 |  |  |

*Note. Anxiety levels were assessed using the Modified Dental Anxiety Scale (MDAS). The comparison between groups was performed with the Mann–Whitney U test (U). No statistically significant differences were found ( $p = 0.232$ ).*

Patient satisfaction, measured using a Likert-type scale based on the Visual Analog Scale (VAS) and administered immediately after the procedure, showed significant differences between the groups. According to **Table 3**, the conventional group had a mean satisfaction score of 13.36 (SD = 2.642), while the digital group had a mean score of 10.78 (SD = 1.458). On this scale, lower scores indicate higher satisfaction. Statistically significant differences were found between the groups (U = 363.5; p < 0.001).

**Table 3.** Comparison of satisfaction levels between conventional and digital impressions.

|  | Mean (M) | Standard deviation (SD) | U | p value |
| --- | --- | --- | --- | --- |
| Conventional | 13.36 | 2.642 | 363.5 | 0.000 |
| Digital | 10.78 | 1.458 |  |  |

*Note. Patient satisfaction was evaluated using a 7-item Likert-type scale based on the Visual Analog Scale (VAS). The comparison between groups was performed with the Mann–Whitney U test (U). Statistically significant differences were found, with greater satisfaction in the digital group ( $p < 0.001$ ).*

## Discussion

This study compared patient-reported dental anxiety and satisfaction between digital and conventional dental impression techniques. While both groups were demographically and socioeconomically comparable at baseline, the findings revealed a significant difference in patient satisfaction, favoring digital impressions, whereas anxiety levels did not differ significantly between groups.

This finding supports existing literature showing that digital impressions are generally better accepted due to increased comfort, reduced gag reflex, and shorter perceived procedure time [4,10,36]. Digital workflows have also been associated with decreased chair time from over an hour to just a few minutes enhancing the overall patient experience [37]. In addition, features such as real-time scanning and the absence of trays may improve patient perception and acceptance [21].

Although our study did not find a significant difference in anxiety, prior research has shown that digital impressions can help reduce anxiety in specific populations, particularly in children, by minimizing intraoral time and discomfort [38]. In adults, however, dental anxiety often depends more on previous experiences, fear of pain, and perceived loss of control than on procedural technique [22]. Communication with the clinician and the clinical environment also play key roles in modulating anxiety [39].

Despite the positive perception of digital impressions, conventional techniques remain necessary in some cases, particularly in full-arch rehabilitations, where accuracy may still favor traditional methods [11]. Moreover, the lack of standardized protocols for digital full-arch impressions highlights the need for further clinical validation [40].

According to numerous studies, digital impressions did not significantly reduce anxiety, they resulted in higher patient satisfaction, supporting their integration into patient-centered prosthodontic care [10,21]. They resulted in slightly lower anxiety levels compared to conventional techniques. In the present study, patients in the digital group reported a mean anxiety score of 8.24 (SD = 3.239), while those who received conventional impressions had a mean score of 8.91 (SD = 3.326). Although the difference was not statistically significant, it reflects a trend consistent with previous findings suggesting that intraoral scanning is better tolerated due to the absence of trays, impression material, and the stimuli that often trigger a gag reflex [3,6,41]. These stimuli have been shown to increase stress in susceptible patients [5,15].

In contrast, satisfaction scores demonstrated a statistically significant difference between groups. Patients who received digital impressions reported higher satisfaction (M = 10.78, SD = 1.458) compared to those in the conventional group (M = 13.36, SD = 2.642), considering that lower scores indicate greater satisfaction. These findings are in agreement with prior studies reporting greater comfort, faster procedures, and reduced retakes with digital workflows [42,43]. Additionally, the perception of modernity and technological sophistication may have contributed positively to the patient experience [24,41].

Although the difference in satisfaction scores between groups was statistically significant, this study was not designed to determine whether the observed difference represents a clinically meaningful improvement from the patient’s perspective. Future research should explore minimal clinically important differences for patient satisfaction measures in dental impression procedures to better contextualize these findings.

While digital impressions offer several benefits in terms of patient comfort and efficiency, the choice of technique should still consider clinical variables such as the extent of the restoration and soft tissue management requirements [13,43]. In edentulous or full-arch cases, limitations in scan accuracy and reproducibility have been noted, supporting the continued relevance of conventional methods in specific scenarios [1,44].

Overall, these results support the integration of digital impressions into routine prosthodontic care, particularly when aiming to improve patient satisfaction [27,45]. At the same time, a patient-centered approach remains essential, as both anxiety and satisfaction are influenced not only by technique but also by communication, trust, and the clinical environment [20,41].

Despite these valuable insights, the study has certain limitations. The sample size, which included 85 participants, provided useful preliminary data but is not sufficient to generalize findings to a larger study sample. Ideally, future research should involve a larger and more diverse sample to strengthen external validity. Another limitation is that the study was conducted within a single faculty, limiting its representativeness across Ecuador. Conducting similar research across multiple faculties nationwide would allow for the identification of regional differences and generate more robust recommendations. Furthermore, the cross-sectional design does not allow for the establishment of causal relationships or the assessment of long-term outcomes.

A potential source of bias was the absence of prior clinical diagnoses of anxiety in some patients. To address this, the Modified Dental Anxiety Scale (MDAS), a validated instrument for objectively assessing anxiety levels in all participants, was used. To maintain consistency in the impression-taking procedures, all operators were postgraduate students in oral rehabilitation from the same academic year, who had completed the same curriculum and clinical training.

This study has limitations inherent to its observational and non-randomized design. Because the impression technique was selected by the treating clinician as part of routine care and was influenced by clinical needs and the patient’s financial means, residual confounding factors and selection bias cannot be ruled out. In particular, socioeconomic factors related to the ability to pay for digital scanning could be associated with patient-reported satisfaction, regardless of the technique itself. Furthermore, the type and complexity of prosthetic treatment were not recorded; therefore, differences in the underlying treatment indication between the groups could have influenced the patient’s experience. Future studies should include treatment characteristics and perform adjusted analyses to better address confounding factors. To address these limitations, future studies should work with a larger sample size and extend the research to several faculties across Ecuador. This would help get results that represent the whole population and allow for the analysis of regional differences. Additionally, since the cross-sectional design limits the ability to establish causal relationships and assess long-term outcomes, future studies should adopt a longitudinal approach to follow patients over time. This would provide a better understanding of how anxiety and satisfaction evolve with repeated exposure to digital or conventional impression techniques, offering deeper insights into patient experiences and treatment effectiveness.

## Conclusion

This study successfully met its objective of comparing anxiety and satisfaction levels in patients undergoing digital versus conventional dental impressions. While no statistically significant differences were observed in anxiety levels between the two methods, patients who received digital impressions reported a higher degree of satisfaction. This suggests that although both techniques may generate similar emotional responses in terms of anxiety, digital impressions are associated with a more comfortable and positively perceived experience.

These results highlight the importance of considering how clinical procedures affect patient comfort and overall perception. In a dental environment where patient-centered care is increasingly prioritized, the adoption of digital technologies can offer advantages that go beyond technical precision. Improved patient satisfaction may lead to better communication, greater trust in the provider, and a higher likelihood of treatment adherence and follow-up attendance.

Moreover, the findings of this study reinforce the value of evaluating both clinical outcomes and the emotional experience of patients. Understanding the psychological impact of procedures allows clinicians to make more informed choices about the techniques they offer. These conclusions also provide a solid foundation for future research, especially studies focused on long-term patient perceptions, behavioral responses to treatment, and the broader integration of digital workflows in routine dental care.

## Ethical Considerations

This study was reviewed and approved by the Comité de Ética de Investigación en Seres Humanos de la Universidad Espíritu Santo (CEISH - UEES) (see S1 File). The study involved only the application of two validated surveys: the Modified Dental Anxiety Scale (MDAS) and a Visual Analog Scale (VAS) for patient satisfaction. Participation was entirely voluntary, and each participant provided informed consent prior to completing the surveys. The consent process was integrated within the digital questionnaire, which was administered via Google Forms. All collected data were anonymized and handled confidentially. The responses were stored in a secure database, accessible only to the members of the research team.

## Data Availability

All relevant data are within the manuscript and its Supporting Information files.

## Financial support

No external funding was received for this study.

## Dissemination plans

The database to corroborate the data shown in this study is available in Supplementary File 4. The data are provided in anonymized form to safeguard the confidentiality of the information provided by the participants in this research.

## Acknowledgements

We would like to express our profound gratitude to all the patients who voluntarily participated in this study, and to the teaching and administrative staff of the Postgraduate Clinic of the Oral Rehabilitation and Implant-Assisted Prosthodontics Program at Universidad de Especialidades Espíritu Santo, for their valuable collaboration throughout the development of this research.

We extend special thanks to our advisor, Dr. Marcelo Armijos Briones, for his constant support and academic guidance throughout the entire process. We also express our gratitude to Dr. Marco Faytong, who, through the final course, provided essential academic foundations for the completion of this work.

Likewise, we express our heartfelt appreciation to our families and friends for their unconditional support throughout this long journey, always guiding us and being our refuge in our most difficult moments. We also thank our dogs, who, with their quiet companionship and boundless love, offered us comfort and joy during the most intense days.

And above all, we thank God for giving us health, clarity, and strength to complete this stage with commitment and gratitude.

## Supporting information

**S1 File. Ethics approval document (CEISH-UEES).**

This file contains the official ethics approval issued by the Comité de Ética de Investigación en Seres Humanos (CEISH-UEES).

**S2 File. Modified Dental Anxiety Scale (MDAS).**

This file contains the MDAS questionnaire instrument used to assess dental anxiety levels.

**S3 File. Visual Analog Scale (VAS) questionnaire for patient satisfaction.**

This file contains the validated VAS-based questionnaire administered after the impression procedure.

**S4 File. Primary Database**

IBM SPSS version 25 file.

## Notes

Conflict of interest: The authors declare that they have no conflict of interest during the development of this research protocol.

### Competing Interest Statement

The authors have declared no competing interest.

### Funding Statement

The author(s) received no specific funding for this work.

### Author Declarations

This research was conducted with the approval of the Ethics Committee for Research in Human Subjects of the UEES (CEISH UEES), which is authorized by the Ministry of Public Health of Ecuador to approve research. The approval code is C-UEES-25-01. Informed consent was obtained digitally via Google Form.

